# Deep phenotyping of women with endometriosis-associated pain and bladder pain syndrome: the TRiPP (Translational Research in Pelvic Pain) study protocol

**DOI:** 10.1101/2022.05.16.22274828

**Authors:** Lysia Demetriou, Lydia Coxon, Michal Krassowski, Nilufer Rahmioglu, Lars Arendt-Nielsen, Qasim Aziz, Christian M. Becker, Judy Birch, Franscisco Cruz, Anja Hoffman, Andrew W. Horne, Lone Hummelshoj, Stephen McMahon, Jane Meijlink, Esther Pogatzki-Zahn, Christine B. Sieberg, Irene Tracey, Rolf-Detlef Treede, Stacey A. Missmer, Krina T. Zondervan, Jens Nagel, Katy Vincent

**Author notes:** Joint first authors (Lysia Demetriou, Lydia Coxon, Michal Krassowski). Deceased (Stephen McMahon).

## Abstract

**Objectives:** Chronic pelvic pain is common, poorly understood, and many women suffer for years without proper diagnosis and effective treatment. The Translational Research in Pelvic Pain (TRiPP) project takes a phenotyping approach, with a particular focus on endometriosis-associated pain (EAP) and bladder pain syndrome (IC/BPS), to improve our fundamental understanding of chronic pelvic pain. We believe that reconceptualising these conditions in the context of the multisystem dysfunction known for other chronic pain conditions rather than as end-organ pathologies has the potential to improve our understanding of the conditions. Our approach combines clinical, biological, physiological and psychological data to establish perturbations in the functions of pain-relevant systems that are specific to EAP and IC/BPS, and those that overlap both conditions and chronic pelvic pain more generally and associated quantitative biomarker profiles.

**Discussion:** We believe that TRiPP’s novel methodological approach will produce clinical data to aid our understanding of pelvic pain and identify underlying pathways for the development of refined animal models and targeted therapeutic treatments.

## Introduction

Chronic pelvic pain (CPP) is common, affecting 5-26.6% of women (1–4). It has significant impact on quality of life and high associated costs for individuals, society and healthcare providers (5). Yet, CPP is neglected and underfunded in both pain and women’s health research fields.

The Translational Research in Pelvic Pain (TRiPP) project (https://www.imi-paincare.eu/PROJECT/TRIPP/) is a collaboration between academics in the UK, Europe and the USA, European Pharma companies, a small-medium sized enterprise (SME) and patient partners. It focusses on endometriosis-associated pain (EAP) and interstitial cystitis/bladder pain syndrome (IC/BPS) in women, two conditions where CPP is highly prevalent and for which current treatments predominantly target the periphery (ectopic tissue for endometriosis and the bladder for IC/BPS) and are often ineffective (6). Our hypothesis is that the pain symptoms in these conditions are generated and maintained by mechanisms similar to those found in other chronic pain conditions but occur in combination with specific pathological lesions and symptoms (8,9). We believe that reconceptualising these conditions in the context of the multisystem dysfunction known for other chronic pain conditions rather than as solely end-organ pathologies has the potential to improve our understanding of the conditions, allow us to identify meaningful subgroups of patients, develop better preclinical models and thus ultimately facilitate research and drug development in this field. The present project takes a deep-phenotyping approach, combining clinical, biological, physiological and psychological data, to improve our understanding of CPP in women.

### Aims/Objectives

The primary objective is to establish perturbations in the function of pain-relevant systems/mechanisms that are specific to EAP and IC/BPS, and those that overlap both conditions (EABP) and CPP more generally. The secondary objectives are to: a) establish biomarker profiles that are specific to EAP and IC/BPS and those that overlap both conditions and CPP more generally; b) determine whether women with pelvic pain can be stratified based on clinical data, the function of pain-relevant systems and their biomarker profile and identify any key pathways/processes underlying pain in these subgroups that may point to novel therapeutic targets; and c) explore whether any subgroups identified during the study relate to response to previous treatment.

## Study Design

TRiPP is an observational cohort study being conducted in phases and includes five groups of participants: EAP: endometriosis-associated pain; EABP: endometriosis-associated and bladder pain; BPS: interstitial cystitis/bladder pain syndrome; PP: pelvic pain with no endometriosis or bladder symptoms; and CON: controls with no pelvic pain or bladder symptoms; (Figure1). The study builds upon two existing endometriosis cohort studies in Oxford and Boston (EndOX: A study to identify possible biomarkers in women with endometriosis, Oxford REC ref 09/H0604/58; Boston Center for Endometriosis (BCE): A Cross-Institutional Biorepository and Database, IRB-P00004267). Given that neither cohort has a specific focus on bladder pain we planned to recruit additional women with IC/BPS from Instituto de Biologia Molecular e Celular (IBMC) Porto. Overall we aimed to identify 800 women with complete baseline questionnaires and available biospecimens (collected according to Endometriosis Phenome and Biobanking Harmonisation Project (EPHect) recommendations) (10,11) who meet our inclusion/exclusion criteria to form the TRiPP cohort.

**Figure 1.**
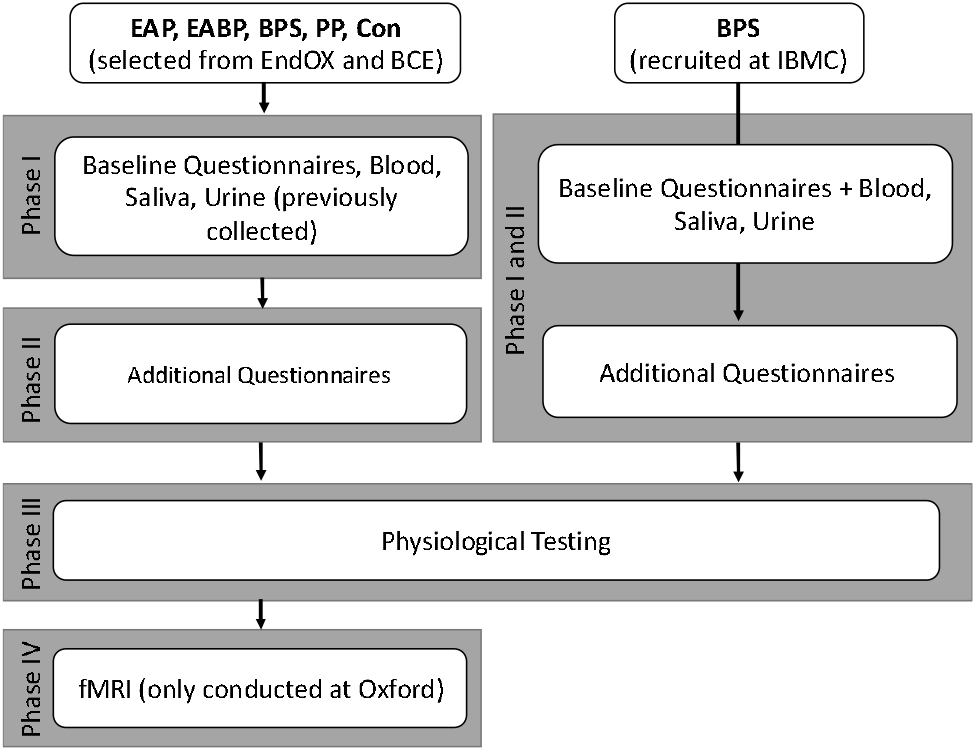
TRiPP Study Design. Phase I in Oxford and Boston conducted prior to TRiPP commencement. Participants from Oxford and Boston selected based on inclusion criteria from available cohorts (EndOX and BCE). Phase I recruitment at Porto (IBMC) commenced with study. Phase II included additional questionnaires completed online or on paper for all sites. Phase III carried out by trained researchers at each site. Phase IV carried out at Oxford by trained researchers.

From this cohort we aimed to recruit 200 participants to undergo psychophysiological testing and of these, 100 who would also undergo a neuroimaging scan.

Data from each study phase will be analysed separately initially, with subsequent multi-modal analysis integrating the different datasets.

Data collection is conducted at the University of Oxford, Boston Childrens Hospital, Brigham and Women’s Hospital and IBMC Porto. The study commenced in April 2018 and is still ongoing, with TRiPP specific recruitment having commenced in September 2019 after all regulatory approvals were gained. All recruitment is now complete.

### Study Population and sample size

Participants forming the TRiPP cohort were selected according to the following criteria: All groups:

- Aged 18 – 50 years (acknowledging some will have been younger when they first contributed data to ENDOx/BCE)
- Not pregnant, lactating or planning pregnancy during the course of the study

EAP:

- surgical diagnosis of endometriosis;
- at least one pelvic pain score of >4/10 on numerical ratings scales (NRS)
- no urinary symptoms (e.g. urge, frequency)
- pain not perceived as arising from the bladder.

EABP:

- surgical diagnosis of endometriosis;
- at least one pelvic pain score of >4/10 on NRS
- urinary symptoms
- pain perceived as arising from the bladder and from other area(s) of the pelvis.

BPS:

- no history of endometriosis;
- at least one pelvic pain score of >4/10 on NRS
- urinary symptoms
- pain perceived as arising from the bladder.

PP:

- no history of endometriosis;
- at least one pelvic pain score of >4/10 on NRS
- no urinary symptoms
- pain not perceived as arising from the bladder.

CON:

- no history of endometriosis;
- no pelvic pain, all scores <3/10 on NRS
- no urinary symptoms

Regretfully, due to the COVID-19 pandemic recruitment of new IC/BPS participants had to be halted in March 2020 and therefore the majority of those in the BPS group were also identified from the existing studies. The full TRiPP cohort therefore comprises 786 women divided into the five study groups. Participants who agreed to participate in TRiPP specific phases of the study could choose which part(s) of the study they wished to be involved in as long as they are able to give informed consent for their participation. For phase IV (fMRI) they need to meet appropriate inclusion/exclusion criteria regarding MRI magnet safety.

The sample size of the omics biomarker discovery analyses in the main cohort was chosen to maximise power of discovery within the most easily back translatable biospecimen (blood), allowing selection of promising, robust signals that can be targeted for further analysis as per existing literature (12). The sample size for the subsequent phases of the study were determined on the basis of previous literature, however, a pragmatic decision about closing recruitment was required in the light of the COVID-19 pandemic to allow time to complete all analyses.

Our full cohort comprises 786 participants, with 158 having completed additional questionnaires and 85 the physiological testing paradigm. fMRI data collection is ongoing.

### Outcome Measures

#### Baseline Questionnaires

All women identified from ENDOx and BCE had already completed the baseline questionnaire (10) and these data have been harmonised in a REDCap database. This questionnaire provides a comprehensive profile of the participants by including gynaecological, obstetric and medical history; detailed assessment of their pelvic pain now and previously; and important demographic variables. Participants recruited at IBMC completed the baseline and additional questionnaires once consented and these data were entered in the database. Women recruited to phase II from Oxford and BCH completed additional questionnaires on paper or directly into the database. Data entered by hand from paper versions was double entered to reduce error for a minimum of ∼20% of the questions for each participant. All questionnaires were available in English and Portuguese. For the Portuguese version, validated translations were used where available, otherwise forward and back translation was used.

### Additional Questionnaire Development

The questionnaires for Phase II of the study were selected by the TRiPP consortium with additional input from members of IMI-PainCare (EPZ, RDT). They were chosen to allow a wide understanding of CPP in the context of what is known about other chronic pain conditions (e.g.mood, catastrophizing, sleep disturbances) (13–15). Some of these domains have been assessed previously in endometriosis and/or IC/BPS (e.g. gastrointestinal symptoms) but not in combination with the variety of other measures collected here, whilst other areas have not been assessed before (e.g. a detailed characterisation of flares in endometriosis). Table 2 illustrates the domains being assessed and tools that are used for phase II.

### Physiological assessments

Very little is known about the neurophysiology of women with CPP. The Multidisciplinary Approach to the Study of Chronic Pelvic Pain (MAPP) Network has undertaken many of these tests in those with Urologic Chronic Pelvic Pain Syndrome (an overarching term that includes IC/BPS), however their cohorts comprise both men and women (https://www.mappnetwork.org). Data from this group and others looking at women only do support the idea that these conditions are similar to other chronic pain conditions (16,17); however, to date this has not been comprehensively assessed by combining multiple assessments in a deeply phenotyped cohort and in combination with endometriosis. The paradigm for TRiPP was designed with input from pain researchers within the TRiPP consortium (LAN, QA, SM, CS, KV) and IMI-PainCare project (EPZ, RDT). We aim to assess all relevant systems whilst keeping the paradigm an acceptable length and avoiding invasive procedures. Patient partners (JB, LH, JM) were key to designing an acceptable paradigm. In order to ensure reproducibility between the centres, all researchers involved in the physiological data collection attended a coordinated training session in Heidelberg, Germany in September 2019 and subsequent post-COVID virtual refresher sessions. The components of the paradigm are listed in Table 2. A questionnaire assessing state measures considered important covariates for the analysis of these physiological data is completed prior to each experimental session (“How are you today?” questionnaire).

### Omics

EDTA plasma from biobanks (stored for a maximum of 8 years) and newly recruited patients was used to obtain genotypes of participants and quantify the relative abundance of proteins and metabolites. 182 proteins were measured using Olink’s proximity extension assay inflammation and neurology panels (18) and ∼1200 metabolites were quantified using ultra-high performance liquid chromatography-tandem mass spectrometry run in four different modes by Metabolon.

### fMRI

Neuroimaging will be used to further understand the central processes underlying observations from the physiological testing and explore whether MRI markers observed in other chronic pain conditions are present in CPP. The scanning protocol includes structural and functional sequences (see Table 1) and is aligned to the fMRI protocol of the BioPain subproject of IMI-PainCare. Punctate stimuli will be used for the “pain task” as they have been previously demonstrated to give robust activation of key pain-related areas in hyperalgesic states (both experimental (19–21) and clinical (22)) and in women with endometriosis-associated pain without hyperalgesia (KV unpublished data).

**Table 1.**
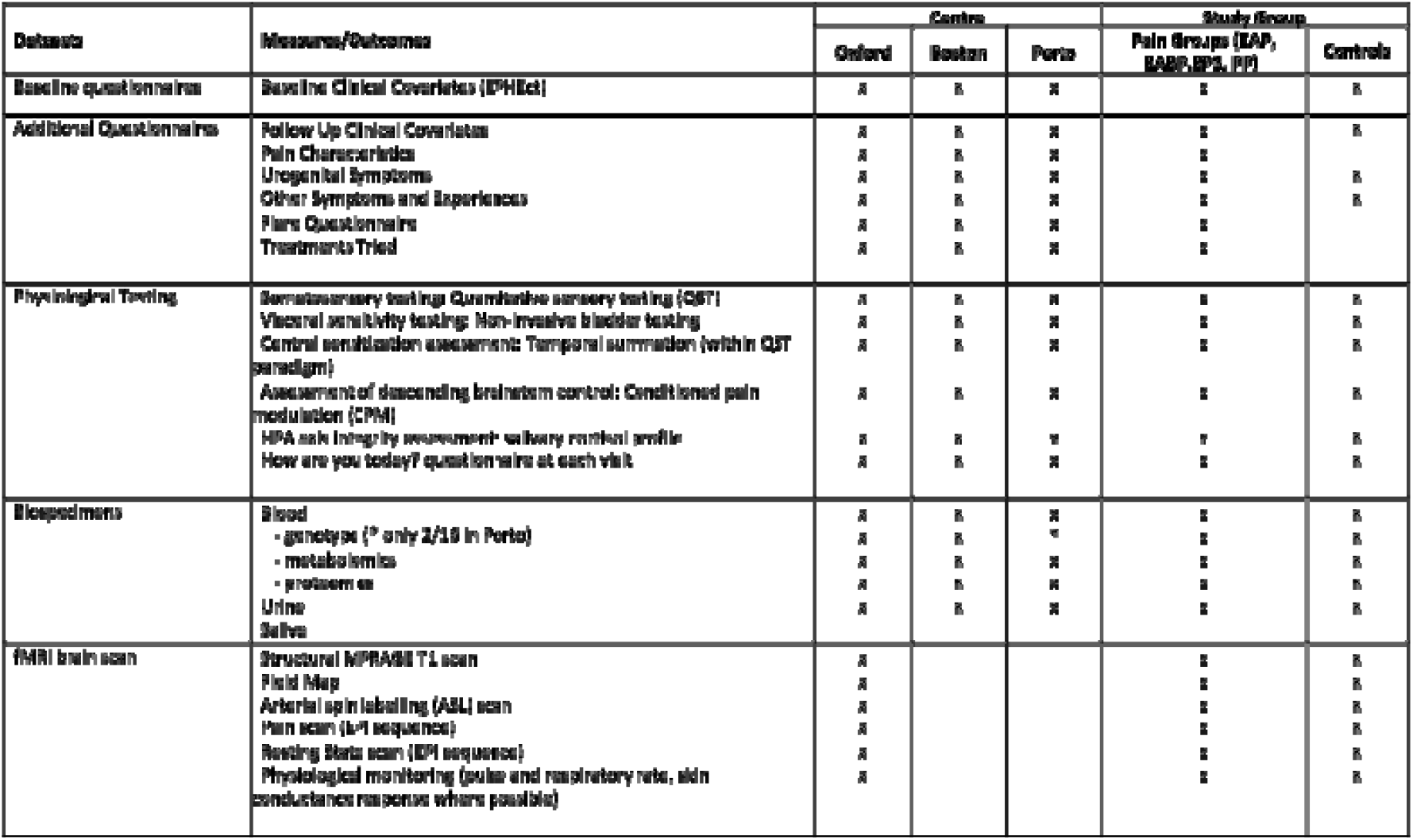
TRiPP Study Measures and Outcomes.

**Table 2.**
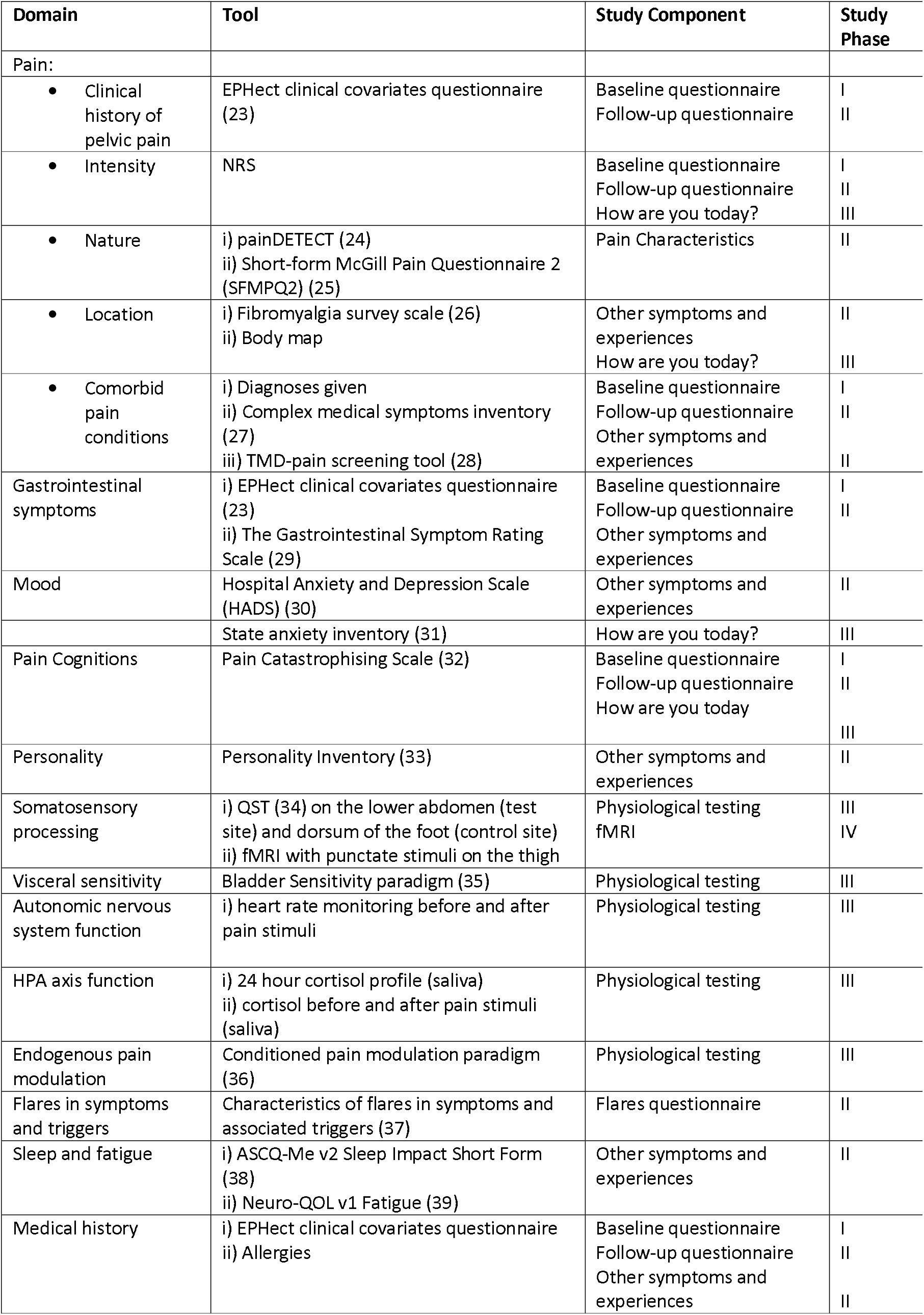
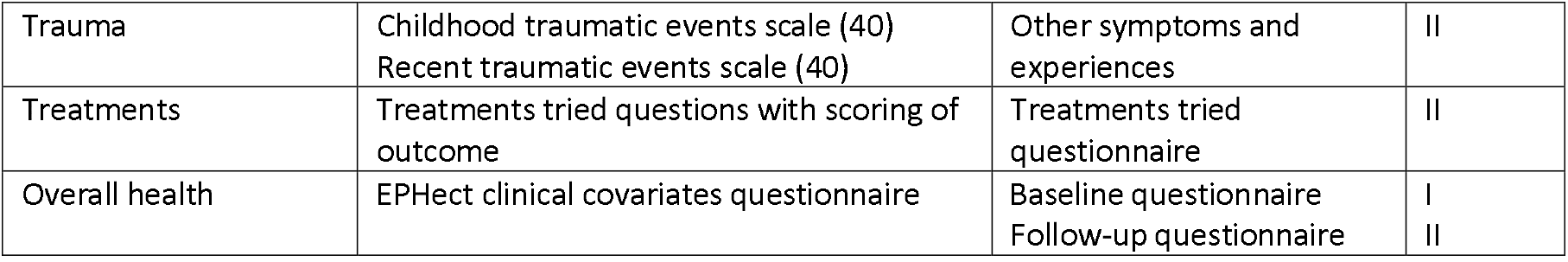
Study Paradigm. Assessment-tools employed at each study phase based on the domains of interest.

### Data Management and Analysis

Participants are only identified by study specific participant number in any database. Personal information is stored separately from these databases and retained at the individual sites. Physiological data was transferred to the University of Oxford prior to analysis.

Derived variables will be calculated from all validated scales and physiological testing paradigms according to published methodology. Regular meetings of the pain data and the omics data analysis groups review all analysis plans before commencing analysis and the overall TRiPP statistical analysis plan is maintained as a live document. Appropriate statistical software will be used to analyse the large volumes of data collected. Omics modelling will be performed using base R, rms and easystats packages, Stan or PyMC3, and multiple Python packages including pingouin. QST data will be analysed in MATLAB. fMRI data will be analysed with FSL (FMRIB’s Software Library; www.fmrib.ox.ac.uk/fsl). Other statistical analysis will be carried out in SPSS. An integration analysis will be undertaken to combine all the different modalities, allowing comparisons of the biomarker profiles between patient groups and controls and an exploration of different phenotypic presentations.

## Discussion

The TRiPP consortium benefits from global interdisciplinary co-operations between clinicians and basic scientists from a wide variety of backgrounds working both in academia and the pharmaceutical industry, as well as three active patient organisations who have been involved with the study since inception. We believe that our novel approach, both in terms of deep phenotyping and the use of methodologies not usually applied to these conditions, has the potential to improve our understanding of pelvic pain. We hope to be able to produce novel clinically relevant data (e.g. an understanding of the prevalence and impact of both widespread pain and flares in CPP), identify underlying pain pathways and thus potential new therapeutic targets or opportunities to repurpose treatments from other conditions. Moreover, other work-packages within the project will back-translate our findings to refine existing rodent models (41) of endometriosis and IC/BPS with the hope of improving the drug development capabilities.

## Limitations

The use of existing bioresources, whilst allowing this project to proceed relatively rapidly, means that the time between biospecimen collection and analysis varied for each participant and resulted in sample aging of relevance to the omics data. Common to most recent studies, the COVID-19 pandemic and related restrictions severely delayed recruitment and data collection for phases III and IV. Additionally, it limited our ability to recruit a new IC/BPS cohort of adequate size. Whilst we could mitigate against this to an extent by using participants within the bioresources who met the BPS recruitment criteria, this group is not of the size we had originally planned and the majority of participants were identified from gynaecology rather than urology clinics.

## Data Availability

All data produced in the present study are available upon reasonable request to the authors

## Declarations

### Ethics approval and consent to participate

The study has received ethical approval from Yorkshire & The Humber – South Yorkshire Research Ethics Committee (19/YH/0030) with local site approvals and is being conducted in accordance with the principles of the Declaration of Helsinki and with relevant regulations and Good Clinical Practice. Informed consent is obtained from all the participants at each phase of the study, and they were informed that they are free to withdraw from the study at any time.

### Availability of data and material

Direct access will be granted to authorised representatives from the Sponsor and host institution for monitoring and/or audit of the study to ensure compliance with regulations. All study data will be stored for 10 years after the end of the study. Once the study and all follow up analyses are complete de-identified data will be deposited in a publicly accessible repository as required by the funders.

### Funding

This project has received funding from the Innovative Medicines Initiative 2 Joint Undertaking under grant agreement No [777500]. This Joint Undertaking receives support from the European Union’s Horizon 2020 research and innovation programme and EFPIA. Further information is found under www.imi-paincare.eu and www.imi.europa.eu.

#### Disclaimer

The statements and opinions presented here reflect the author’s view and neither IMI nor the European Union, EFPIA, or any Associated Partners are responsible for any use that may be made of the information contained therein.

## Acknowledgements

The authors wish to acknowledge Allison Vitonis and Kurtis Garbutt for their work in harmonising databases between Boston and Oxford and identifying participants suitable for the TRiPP cohort.

## Consent for publication

All authors read and approved the final manuscript.

## Competing interests

LD, LC and MK declare no competing interests.

NR: No competing interests

LAN: No competing interests

QA: No competing interests

CMB: Research Grants from Bayer Healthcare, MDNA Life Sciences, Roche Diagnostics, European Commission, NIH. His employer has received consulancy fees from Myovant and ObsEva for work outside of this project.

JB: No competing interests

FC: Consultant and/or investigator for Allergan (Abbvie), Astellas, Bayer, Ipsen and Recordati

AWH: reports grant funding from the MRC, NIHR, CSO, Wellbeing of Women, Roche Diagnostics, Astra Zeneca, Ferring, Charles Wolfson Charitable Trust and Standard Life. His employer has received consultancy fees from Roche Diagnostics, AbbVie, Nordic Pharma and Ferring, outside the submitted work. In addition, AWH has a patent for a serum biomarker for endometriosis pending.

LH: no competing interests in relation to this work

JM: no competing interests

EMP-Z: received financial support from Grunenthal and Mundipharma for research activities and advisory and lecture fees from Grünenthal, Novartis and Mundipharma. In addition, she receives scientific support from the German Research Foundation (DFG), the Federal Ministry of Education and Research (BMBF), the German Federal Joint Committee (G-BA) and the Innovative Medicines Initiative (IMI) 2 Joint Undertaking under grant agreement No 777500. This Joint Undertaking receives support from the European Union’s Horizon 2020 research and innovation programme and EFPIA. All money went to the institution E.M.P.-Z. is working for.

AH: Employee of Bayer AG, Germany. CS: declares no competing interests

IT: On the scientific advisory board for Amgen (2017–2019); has received honoraria for providing various educational talks and workshops; is a council member of the Medical Research

Council of the United Kingdom; is a trustee of MQ, a mental health charity; is President-elect of FENS; and is involved with Innovative Medicines Initiative–PainCare (Biopain), a pan pharma-academia consortium.

RDT: Ad board for BAYER, IASP task force on chronic pain classification

SAM: has been an advisory board member for AbbVie and Roche and receives research funding from the National Institutes of Health, the US Department of Defense, the J. Willard and Alice S. Marriott Foundation, and AbbVie; none are related to the presented work.

KTZ: reports grant funding from EU Horizon 2020, NIH US, Wellbeing of Women, Bayer AG, Roche Diagnostics, Evotec-Lab282, MDNA Life Sciences, outside the submitted work.

JN: Employee and shareholder of Bayer AG, Germany

KV: declares research funding from Bayer Healthcare and honoraria for consultancy and talks and associated travel expenses from Bayer Healthcare, Grunenthal GmBH, AbbVie and Eli Lilly.

## Author’s contributions

LD, LC, MK and KV drafted the manuscript. All other authors contributed to the original design of the project and reviewed and edited the manuscript.

## Figures and Tables

See our general formatting guidelines. Authors must limit the number of tables and figures in the manuscript to **3** in order to be consistent with a note article type.

Additional tables/figures can be provided as additional files or be deposited in an appropriate data depository (e.g. Figshare).

